# Public-private mix in health systems and repercussions for health inequalities in Latin American countries: a scoping review protocol

**DOI:** 10.1101/2024.06.01.24308311

**Authors:** Eduarda Ferreira dos Anjos, Suelen Carlos de Oliveira, Adriana Mendoza-Ruiz, Maria del Pilar Flores-Quispe, Adelyne Maria Mendes Pereira, Carl Kendall, Ligia Regina Franco Sansigolo Kerr, Naiá Ortelan, Elzo Pereira Pinto Junior, Natalia Romero, Alastair Leyland, Mauricio Lima Barreto, Cristiani Vieira Machado, Unit on the Social and Environmental Determinants of Health Inequalities (SEDHI)

## Abstract

**Introduction:** Health system models in Latin American countries express differences in the state’s role in regulating, financing and providing health services, as well as in the coexistence of different care arrangements involving public and private entities. This scoping review will seek to identify evidence on how the different public-private configurations of health systems influence health inequalities in Latin America.

**Methods and Analysis:** This protocol will be guided by the scoping review methodology developed by the Joanna Briggs Institute. The results will be presented according to the PRISMA-ScR protocol. Searches will be carried out on the Scielo, Lilacs, Embase, Pubmed, Web of Science and Scopus databases. The inclusion criteria will be publications whose central theme is the public-private mix of health systems in Latin America between January 2000 and March 2024. Exclusion criteria will be clinical trials, incomplete studies or in the design phase, duplicate publications, pre-print studies and gray literature (interim reports, unpublished texts, dissertations, and theses. The steps for carrying out the scope review will: (1) identifying the research question; (2) identifying relevant studies; (3) study selection; (4) charting the data; and (5) collating, summarizing, and reporting the results. Two independent reviewers will select the articles. The results will be described, analyzed, and categorized through narrative synthesis, correlating with the research objectives and questions.

**Expected results:** It is hoped that this scoping protocol will provide a comprehensive framework for investigating the gaps in the current literature on the public-private mix in Latin America and the effects on health inequalities.

**Link to the protocol record in the Open Science Framework (OSF):** https://osf.io/rkzx3/?view_only=4594a2c52f2d47128d805fdd9dd1359b.

## Introduction

Health systems in Latin American countries have been structured in various ways throughout the 20th century, generally involving a strand of collective public health actions and another of medical care aimed at workers in the formal labour market. Throughout their history, the consolidation of these systems has been hampered by economic restrictions, democratic instability, social inequality, and widespread informality in the labour market ^1^. The diversity of ethnic and cultural characteristics, the population’s lifestyles (urban, rural, and remote rural) and the socioeconomic level of the population in the different nations of Latin America add to the challenges for health systems ^2^.

Reforms adopted over the years in various regional countries expanded the private health sector under state incentives associated with new business dynamics, resulting in a complex public-private mix ^3, 4, 5^. This complexity is expressed in the segmentation of the population according to their affiliation to services, the composition of the supply of care, health spending, the diversity of providers working in the sector, the multiplicity of health professionals’ ties, as well as the different coverage and access between groups according to their ability to pay or pay directly for services ^6, 7, 2^.

In addition, there are historical-structural challenges in the organization, financing, and provision of health systems in Latin America, which have recently been highlighted by the COVID-19 pandemic, which has exacerbated the inequalities and weaknesses of these systems, revealing their limitations and potential ^8, 9, 10^. In this sense, understanding public-private health arrangements in Latin America over the last few decades and considering the impacts of the recent pandemic can help direct public policies with a focus on state-market relations. Thus, this protocol proposes a scoping review to identify and explore the evidence on the effects of the public-private mix of health systems on health inequalities in Latin America.

## Methods and Analysis

The review will cover the period from 2000 to 2024. The time frame is justified because it covers more than two decades marked by changes in various health systems in Latin America but with persistent health inequalities. The scoping review technique was selected for its potential in mapping complex and heterogeneous literature, considering the particular contexts of Latin American countries, as well as being useful for providing decision-makers with information on how the topic has developed over time, as well as gaps and the need to strengthen public policy agendas.

The construction of this protocol was based on internationally recognized and validated scoping review guidelines, as well as articles in the field of public health that have adopted this methodological design. This review follows the Joanna Briggs Institute (JBI) and the Preferred Reporting Items for Systematic Reviews and Meta-Analyses Extension for Scoping Reviews (PRISMA-ScR) and will follow the following methodological steps: (1) identifying the research question; (2) identifying relevant studies; (3) study selection; (4) charting the data; and (5) collating, summarizing and reporting the results.

This protocol was registered on the Open Science Framework platform following the JBI recommendation, to conduct an impartial review, avoid duplication of research, and guarantee the originality of the work. It can be consulted via the register: https://osf.io/rkzx3/?view_only=4594a2c52f2d47128d805fdd9dd1359b.

### Step 1: Identifying the research question

Public-private relations in health systems and their implications for health inequalities in Latin America are the focus of this review. In particular, the participation and arrangements between the public and private sectors in the financing, coverage, and provision of services will be highlighted. The focus, question and hypothesis of this work are detailed in Box 1.

### Box 1. Focus, question, and hypothesis of this scoping review

The construction of the research questions was based on the PCC mnemonic, where ‘P’ represents the population, ‘C’ the concept and ‘C’ the context. The ‘population’ was taken to be the equivalent of the unit of analysis for public health (in this case, “Health Systems”). For the context, in addition to the term Latin America, seven countries were included in the bibliographic search: Argentina, Brazil, Chile, Colombia, Ecuador, Mexico and Peru (Box 2).

### Box 2. Population (P), Concept (C) and Context (C)

The seven countries were selected because they are populous and concentrate most of the region’s publications, as well as because they meet criteria of interest for the analysis, such as the persistence of significant socioeconomic inequalities and the presence of varied health system arrangements, including the universal model, the social insurance model, and different private sector financing and provision models.

The health systems of the seven countries selected for the study are briefly described in Box 3, covering coverage, regulation, financing and system organization. Of these, three are federal countries (Argentina, Mexico and Brazil) and four are unitary republics (Chile, Colombia, Peru and Ecuador).

### Box 3. General characteristics of Health Systems in seven countries of Latin America

The seven countries show different levels of socioeconomic and health inequalities. The data related to the Gini coefficient reiterates the socioeconomic disparities in Latin American countries. In 2020, the average coefficient was 0.46 for the entire region. Ecuador (0.46) and Peru (0.46) have indicators similar to or close to the regional average, making it clear that they face different gradients of inequalities in terms of health and socioeconomic conditions. Mexico (0.45) showed the smallest difference between the richest and poorest sections of its population. In contrast, Colombia (0.55), Brazil (0.51) and Chile (0.47) stood out for their greater concentration of income, making them among the most unequal countries in the world ^35^.

The Latin American federations, Brazil represents the case of a universal health system, based on a comprehensive conception of social security, with many differences in access to health between regions of the country and a strong presence of the private sector; while Mexico has a social insurance system, with high private spending through direct disbursement by families. The Argentine system has maintained its corporate base, with the presence of the private sector in the provision of services and the existence of specific public programs ^6^.

Chile and Colombia are illustrative cases of neoliberal reforms that radically altered the role of the state and markets in health. In Chile, the reform began at the end of the 1970s under the military dictatorship, resulting in a dual system. In Colombia, in the 1990s, under a democratic regime, the Structured Pluralism model was implemented ^24, 19^. Ecuador and Peru are characterized by mixed systems, with a weakened public sector compared to the private sector ^31, 32, 33, 29, 30, 27^.

### Step 2: Identifying relevant studies

#### Data sources

The search for scientific articles will be carried out in six databases: Lilacs via the Biblioteca Virtual de Saúde (BVS in Portuguese); Embase; Pubmed; Scielo; Web of Science; and Scopus. Access to the Web of Science and Scopus databases will be via the Portal de Periódicos Capes (in Portuguese). In addition, references relevant to the topic will be included in the articles found.

#### Search strategy

The search will be carried out using a main search strategy which will be adapted for each database according to the PCC and the defined inclusion and exclusion criteria. The search strategies for the three databases with controlled vocabulary, Lilacs/BVS, Embase and Pubmed, will use the English language descriptors indexed: Descriptors in Health Sciences (Decs in Portuguese), Emtree term and Medical Subject Headings (MESH), respectively (Box 4). In the other databases, the strategy will exclusively use keywords (Appendix).

### Box 4. Search terms

#### Step 3: Study selection

After searching in the descriptor and/or keyword fields (depending on the database), the texts will first be screened by reading the title and abstract and then by reading the full text.

The selection of the documents to be included in this scope review at each stage will be carried out by two reviewers independently. Any disagreements will be resolved by consensus or by decision of a third reviewer. To increase uniformity and accuracy, all reviewers will first carry out a calibration exercise. Subsequently, reviewers in pairs will independently analyze all the records using the Rayyan platform (https://www.rayyan.ai/). Full articles that meet the inclusion criteria established for the study will be selected and examined in detail.

Monitoring of the study selection process will be detailed in a flowchart (according to PRISMA-ScR guidelines), according to the identification, selection, eligibility, and inclusion stages. The search results will be managed using the ‘Zotero’ software to collect, store and organize the references. Using the reference manager will allow for organization into different groups, facilitating sharing with the researchers participating in this study.

#### Inclusion criteria

This review will consider publications whose central theme is the public-private mix of health systems in Latin America (especially those dealing with Brazil, Ecuador, Mexico, Chile, Colombia, Peru and Argentina, individually or comparatively with other countries), published between January 2000 and March 2024.

The identification and selection of relevant publications will include the following types of documents: essays, qualitative and quantitative research (case studies, ecological, cross-sectional and time series studies), analysis and evaluation of health policies and systems, published in English, Portuguese, and Spanish.

Only open-access scientific articles published in journals indexed in the aforementioned databases will be included, as they undergo peer review. Theses and dissertations will not be included (since their end product is usually scientific articles published in journals) and books or chapters (the databases selected are incomplete for such works, and not all are peer-reviewed and open access).

#### Exclusion criteria

Clinical trials, incomplete studies or in the design phase, duplicate publications, pre-print studies, systematic reviews, narrative reviews and gray literature (interim reports, unpublished texts, dissertations and theses) will be excluded.

### Step 4: Charting the data

The search strategy will be carried out in three stages: (1) Initial search in two databases; (2) Identification of terms to be included, enrichment of the search strategy and search in the other databases; and (3) Review of references of included documents to track down and evaluate the inclusion of other works. This strategy will be guided by a data extraction form developed electronically to extract the information, which will be filled in independently by two researchers and then converted into a file with a format compatible with the Microsoft Excel® program. The file generated will be the database to be used in the subsequent phase of this review and will contain information identifying the study (title, first author, year, objectives, study design, funding) and data related to the research questions (geographical context/countries).

In this way, the following information will be mapped by the reviewers: characteristics and details of the study (title, first author, year, country, objectives and study design) and data related to the research questions (public-private configuration in financing, coverage and provision of services, as well as repercussions for health inequalities and response to covid-19).

### Step 5: Collating, summarizing and reporting the results

A descriptive analysis of the included studies will be carried out using the concepts described in the form used to extract the data and other characteristics relevant to the findings. The content of the findings will be categorized into “financing models”, “service coverage”, “public-private mix in the provision of services” and “repercussions on health inequalities”.

The results will be summarized to return to the research question and the proposed objective. The scoping review by Nyanchoka et al ^36^ will be used as a reference to choose the appropriate resources for presenting the results.

### Ethical considerations

This research does not require ethical approval because the data used are scientific articles published in indexed journals.

### Disseminations of Knowledge

This scoping protocol will provide a comprehensive framework for investigating the gaps in the current literature on the public-private mix in Latin America and the effects on health inequalities, guided by the guidelines recommended by the Joanna Briggs Institute and the PRISMA-ScR protocol. This evaluation will provide tools and evidence to policymakers of the public-private mix in Latin America and the effects on health inequalities. We will disseminate the data in scientific journals, reports, and policy briefings targeting policymakers and civil society.

## Data Availability

No datasets were generated or analysed during the current study. All relevant data from this study will be made available upon study completion.

## Declarations

### Ethics approval and consent to participate

Not applicable.

### Availability of data and materials

Not applicable.

## Acknowledgments

The authors would like to acknowledge the Unit on the Social and Environmental Determinants of Health Inequalities (SEDHI) by funding through research grants.

## Author’s contribution

Anjos EF, Oliveira SC, Pereira AMM and Mendoza-Ruiz A designed the scoping review. Anjos EF, Oliveira SC, Pereira AMM, Mendoza-Ruiz A, Flores-Quispe MDP, Pinto Junior EP and Machado CV built the protocol and method. Anjos EF, Oliveira SC, Pereira AMM, Mendoza-Ruiz A, Flores-Quispe MDP, Kendall BC, Kerr LRFS, Ortelan N, Pinto Junior EP, Romero N, Leyland A, Barreto ML and Machado CV, drafted the manuscript. All authors approved the final version of the manuscript.

